# Effectiveness of screening and ultra-brief alcohol intervention in primary care: a pragmatic cluster randomised controlled trial

**DOI:** 10.1101/2024.12.27.24319613

**Authors:** Ryuhei So, Kazuya Kariyama, Shunsuke Oyamada, Sachio Matsushita, Hiroki Nishimura, Yukio Tezuka, Takashi Sunami, Toshi A. Furukawa, Ethan Sahker, Mitsuhiko Kawaguchi, Haruhiko Kobashi, Sohji Nishina, Yuki Otsuka, Yasushi Tsujimoto, Yoshinori Horie, Hitoshi Yoshiji, Takefumi Yuzuriha, Kazuhiro Nouso

## Abstract

**Objective:** To evaluate the effectiveness of screening and ultra-brief intervention (Ultra-BI) delivered by primary care physicians in less than 1 minute compared to simplified assessment only (SAO) for reducing alcohol consumption among patients with hazardous drinking.

**Design:** Pragmatic, cluster randomised, parallel-group, superiority trial. We used a computer-generated random sequence to allocate clusters. Only participants and personnel who collected participant-reported outcomes remained blinded.

**Setting:** 40 primary care clinics in Japan, which did not provide routine screening and brief intervention for hazardous drinking, treatment, or self-help groups for alcohol dependence.

**Participants:** 1,133 outpatients aged 20-74 years with hazardous drinking (scores of Alcohol Use Disorders Identification Test-Consumption [AUDIT-C] ≥5 for men and ≥4 for women). Patients who were pregnant or suspected of having COVID-19-like symptoms were excluded.

**Interventions:** Clusters were randomised to Ultra-BI (21 clusters, n=531) or SAO (19 clusters, n=602) groups. Ultra-BI comprised screening with AUDIT, brief oral advice, and an alcohol information leaflet. SAO involved only simplified assessment with AUDIT-C.

**Main outcome measures:** The primary outcome was total alcohol consumption in the preceding 4 weeks (TAC) at 24 weeks post-randomisation. Secondary outcomes included TAC at 12 weeks and readiness to change drinking habits at 12 and 24 weeks.

**Results:** At 24 weeks, the difference in TAC between Ultra-BI (1046.9g/4 weeks, 95% confidence interval [CI] 918.3-1175.4) and SAO (1019.0g/4 weeks, 95% CI 893.5-1144.6) groups was 27.8g/4 weeks (95% CI -149.7 to 205.4). Bayes factor analysis (0.08±0.25) strongly supported the null hypothesis for TAC at 24 weeks. Ultra-BI group showed higher readiness to change drinking habits at both 12 (difference 0.30 [95% CI 0.10 to 0.40]; Hedge’s g 0.21 [95% CI 0.10 to 0.33]) and 24 weeks (difference 0.20 [95% CI 0.10 to 0.30]; Hedge’s g 0.16 [95% CI 0.05 to 0.28]).

**Conclusions:** This trial did not support the effectiveness of Ultra-BI for alcohol consumption compared to SAO, but did improve readiness to change compared to SAO. These findings call for developing effective, low-cost interventions in primary care settings.

**Trial registration:** UMIN000051388

**What is already known on this topic:**

- Brief interventions (BIs) for hazardous drinking have been widely recommended in primary care settings, but implementation rates remain low due to various barriers.
- Ultra-brief interventions (Ultra-BIs) have shown mixed results in different settings, with some studies suggesting they can be as effective as longer advice or counselling.
- No randomised controlled trial has directly investigated the effectiveness of Ultra-BIs over assessment-only control in primary care settings.

**What this study adds:**

- This large-scale pragmatic cluster randomised controlled trial did not support the effectiveness of Ultra-BI on alcohol consumption at 12 and 24 weeks compared to simplified assessment only (SAO) in Japanese primary care settings.
- Ultra-BI showed higher readiness to change drinking habits at both 12 and 24 weeks compared to SAO, despite not reducing alcohol consumption.
- These findings challenge current recommendations for screening and brief interventions in primary care and suggest a need for re-evaluation of these practices.

## Introduction

Globally, harmful alcohol use is a major public health concern, contributing to 3 million deaths and 5.1% of the global burden of disease annually [1]. Hazardous drinking, originally defined by a “quantity or pattern of alcohol use that places patients at risk for adverse consequences” [2], is seen in approximately 20% of patients in primary care settings [3,4] and is associated with a wide range of physical, mental, and social harms [5,6]. To address this global health issue, brief interventions (BIs), which consist of screening and short counselling sessions, have been widely recommended as an effective approach to reduce hazardous drinking in primary care settings [7–9].

The SIPS trial, a large-scale cluster RCT, found that the ultra-brief intervention (Ultra-BI) group, which only received a leaflet with feedback on screening results, showed comparable reductions in hazardous drinking to the more time-intensive BI groups. [10,11]. For policymakers and clinicians, the idea of widely implementing shorter, simpler, and cheaper interventions is highly appealing, given the low implementation rate of standard BI due to time constraints [7,9].

However, the available evidence remains inconclusive as to whether both the Ultra-BI and the more time-intensive BI are equally effective or equally ineffective compared with assessment-only control [12]. Randomised controlled trials (RCTs) comparing Ultra-BI to assessment-only controls have demonstrated mixed results across different settings. Ultra-BI is shorter than standard BI and may only include leaflets, booklets, or computer-generated feedback letters (Cunningham et al. 2012). For inpatient settings, Ultra-BI shows a greater reduction in alcohol consumption compared to assessment-only controls and similar effects to 20-35 minutes of counselling [13,14]. Additionally, our small quasi-experimental study showed promising results suggesting the potential effectiveness of Ultra-BI for alcohol consumption relative to assessment-only control [15]. Conversely, in community and emergency settings, RCTs do not support the effectiveness of Ultra-BI on alcohol consumption compared to assessment-only controls [16,17]. No RCT has directly investigated the effectiveness of Ultra-BI over assessment-only control in primary care settings.

We therefore designed and conducted a large-scale pragmatic cluster RCT in Japanese primary care settings. We aimed to evaluate the effectiveness of screening and the Ultra-BI delivered by primary care physicians in less than 1 minute compared to a simplified assessment only (SAO) control at individual participant level in reducing alcohol consumption at 12- and 24-week follow-up.

## Methods

This was a pragmatic, cluster randomised, parallel-group, superiority trial entitled the EASY (Education on Alcohol after Screening to Yield moderated drinking) study. The main aim of this paper is to report the results of the cluster RCT, which includes intervention after screening and follow-up, in accordance with the CONSORT 2010 extension for cluster RCT [18]. Before initiating the recruitment of screening participants, we prospectively registered the study with the UMIN-CTR (https://www.umin.ac.jp/ctr/) on 23 June, 2023, under the UMIN identifier UMIN000051388. We have reported on the prevalence of hazardous drinking among all participants responding to the screening survey elsewhere [4].

### Settings and procedures

Figure 1 presents the summary of the trial procedures to clarify the sequence and contents of screening, intervention, and follow-up. We invited primary care clinics through personal connections with the authors and local medical collaborations in daily practices. When inviting clinics, we provided videos which introduced the study procedure and demonstration of the Ultra-BI.

**Figure 1.**
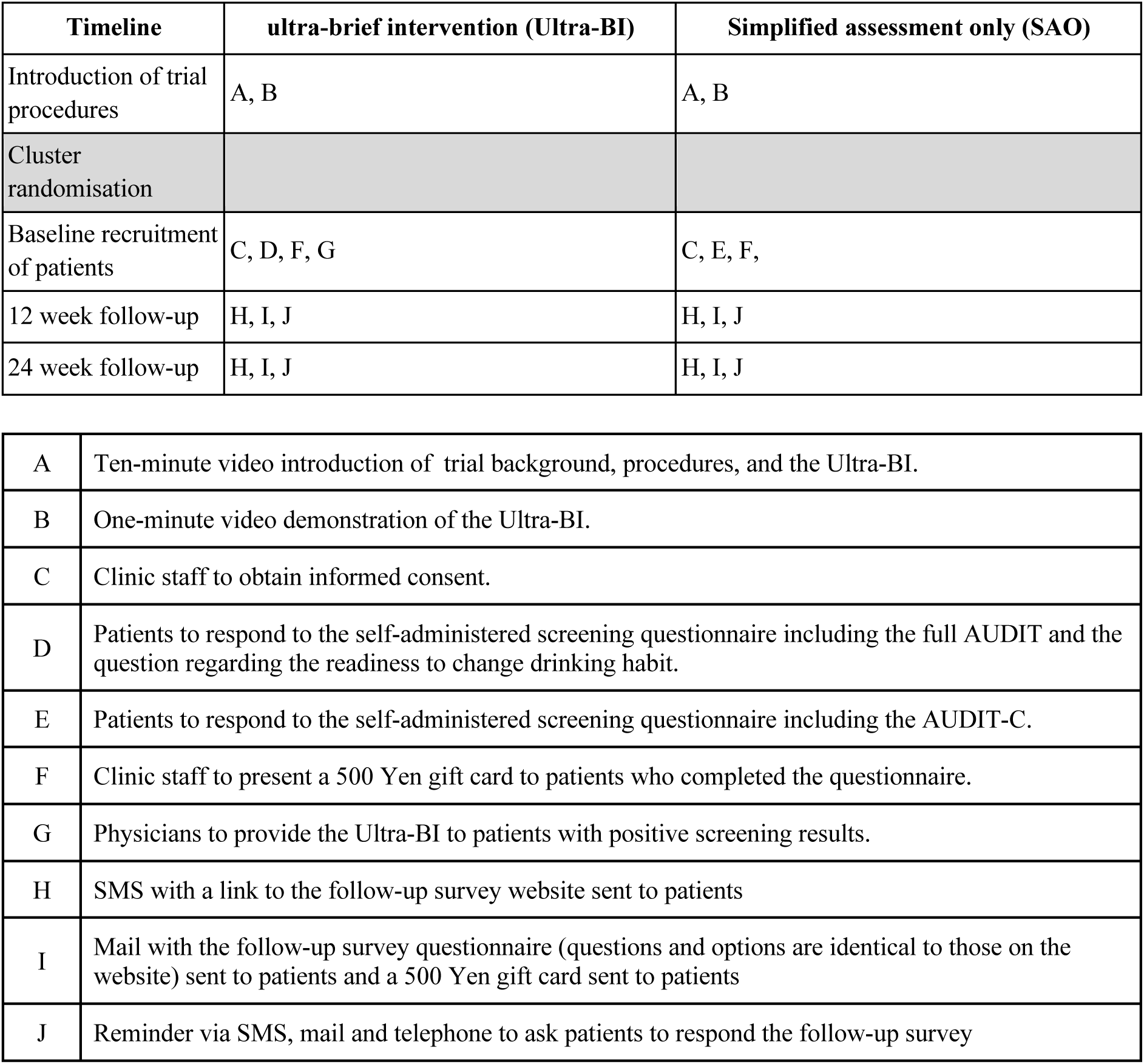
PaT plot of procedures through the trial.

We allocated clusters to the two arms of the study using a block randomisation method without any stratification or matching. The statistician (S.O.), who was not involved in the recruitment of clusters, generated a random sequence on a computer. Another researcher (S.M.), also uninvolved in the recruitment process of clusters, performed the cluster allocation using this random sequence before each site initiated the screening and intervention.

The screening and intervention for participants took place at each cluster between 29 June and 7 August, 2023. We asked each cluster to invite all eligible patients consecutively, until exceeding the target enrolment of 29 patients per cluster. Patients eligible for screening were invited at the clinic reception to provide written consent for the trial participation after reading the informed consent document. To keep participants blinded, the document mentioned possible brief advice from physicians but did not disclose the random allocation or intervention of interest. To replicate real-world clinical settings as closely as possible, no formal oral explanation of the trial was given. The consenting participants responded to the screening survey questionnaire. Those who met the criterion for intervention and follow-up in the Ultra-BI group received the intervention right after screening whereas those in the SAO group did not. Receptionists and physicians remained unblinded throughout this procedure.

We conducted follow-up surveys at 12 and 24 weeks after the screening and intervention to assess the outcomes. Participants were given the option to respond to the follow-up survey questionnaires either in paper form or online. The follow-up survey questionnaires and QR codes for accessing the online survey were sent via postal mail. Additionally, text links to the online survey were sent through mobile phone short messages to maximise response rates. This dual approach ensured that participants could choose the most convenient method for completing the follow-up surveys.

For the 12-week survey, we initially used registered mail requiring signature upon delivery; however, this resulted in some delivery failures due to recipient absence. Therefore, we switched to regular postal mail for the 24-week survey. The mobile phone messages also included a notification about the mailed questionnaires.

The survey company, Neo marketing, Inc., contracted by the research team conducted the administration of follow-up surveys, and the entry of screening and follow-up survey data. If there was missing data, staff of the survey company tried to contact participants via text messages or phone calls to obtain data. The staff members responsible for missing data collection remained blinded to the allocation of trial participants.

### Eligibility criteria for participants and clusters

Inclusion criteria for clusters were: 1) primary care clinics in Japan, and 2) did not previously provide screening with the Alcohol Use Disorders Identification Test-Consumption (AUDIT-C) [19] and BI for hazardous drinking in routine practices, specialised treatment for alcohol dependence, or opportunities for self-help groups focused on alcohol dependence.

Inclusion criteria for participants for screening were: 1) outpatients, 2) age 20 to 74 years at the time of consent, and 3) able to understand the informed consent form and provide written consent. Exclusion criteria for participants at screening were: 1) pregnant women, 2) patients suspected of having COVID-19-like symptoms, and 3) patients judged unsuitable for the study by the physicians at the clusters. An inclusion criterion for the follow-up was hazardous drinking defined as AUDIT-C (AUDIT-C) scores ≥ 5 for men and ≥ 4 for women at the screening survey.

### Measures

#### Screening survey

From all screening participants both at the Ultra-BI and the SAO clinics, we obtained data regarding their age groups, sex, medical histories, types of visits (such as first visit, visit as needed, or routine appointment), and their readiness to change their diet and smoking habits. We included these assessments of diet and smoking habits to mask the study hypotheses related to drinking habits. To assess participants’ readiness to change their diet and smoking habits, we used questions from the Japanese National Health and Nutrition Survey [20]. The response options were combined and reduced to reduce the burden on respondents. To identify participants engaging in hazardous drinking, we used the AUDIT-C (score range 0–12), which comprises questions 1 to 3 of the Alcohol Use Disorders Identification Test (AUDIT) [19] and has been validated for screening use in primary care settings [21]. A validation study in Japan found that a score of ≥ 4 for women and ≥ 5 for men on the AUDIT-C corresponds to a score of ≥ 8 on the AUDIT, indicating hazardous drinking or potential alcohol dependence [19,22]. We calculated the baseline total alcohol consumption in grams per 4 weeks based on the first and second items of the AUDIT-C questionnaire.

Additionally, for the questionnaire given to screening participants at the Ultra-BI clusters, we included questions about readiness to change drinking habits, along with questions 4 to 10 of the AUDIT. We evaluated the readiness to change drinking habits with the question, “Are you considering improving your drinking habits?”, with the following response options: “Already working on improvement“, “Intending to improve”, “Interested but no intention to improve”, “No intention to improve”, and “No improvement needed”. We developed these question and response options based on the Japanese National Health and Nutrition Survey [20], and their psychometric reliability and validity has not yet been confirmed. We did not include the assessment of readiness to change drinking habits in the SAO cluster questionnaires, as the assessment itself might have an effect on behaviour change, potentially making it more difficult to detect the effects of Ultra-BI [23,24].

After the screening survey period, we gathered information from the clusters on the approximate percentages of all potential participants who were asked to participate in the EASY study and agreed to participate. The details regarding these percentages were reported elsewhere [4].

#### Follow-up survey

At 12 and 24 weeks after the screening survey, we obtained data regarding the amount of alcohol consumption and readiness to change diet, smoking, and drinking habits from all trial participants at clusters assigned to both groups. To assess the amount of alcohol consumption, participants were asked about their usual drinking habits, including the type of alcoholic beverages consumed, the types of containers (e.g., cans or glasses), the number of glasses, bottles, or cans, and the frequency of drinking days per 4 weeks.

In terms of readiness to change health-related lifestyles, the questionnaires were the same as those used at the screening survey. However, for the question regarding readiness to change drinking behaviour, the option, “Interested but no intention to improve” was inadvertently omitted from the follow-up survey.

#### Outcomes

The primary outcome was past 4 weeks total alcohol consumption (TAC) at 24 weeks. Using the detailed responses to the follow-up survey questionnaires, we calculated the TAC by referring to a table listing the average alcohol content by volume (ABV) for various types of beverages and the standard volumes of different containers. The TAC was calculated using the following formula: TAC [g/4 weeks] = Volume [ml] × ABV/100 × the number of glasses, bottles, or cans × 0.8 × frequency of drinking days. The initial version of this table was created before the follow-up survey was conducted. If participant responses included beverages or containers not listed in the table, the ABV and standard volume were defined without considering participants’ allocated group.

Additionally, WHO drinking risk level (DRL) was calculated based on TAC [25]. The WHO DRL categorises drinking patterns into different risk levels based on TAC per day. Low risk is defined as up to 40 grams per day for men and up to 20 grams per day for women. Medium risk is 41-60 grams per day for men and 21-40 grams per day for women. High risk is 61-100 grams per day for men and 41-60 grams per day for women. Very high risk is over 100 grams per day for men and over 60 grams per day for women. These thresholds signify the potential health risks associated with alcohol consumption and provide guidelines for reducing such risks.

The secondary outcomes included TAC at 12 weeks, as well as readiness to change drinking habits at both 12 and 24 weeks. For readiness to change drinking habits, “Interested but no intention to improve” and “No intention to improve” were combined into “No intention to improve” due to the inadvertent omission of “Interested but no intention to improve” from the follow-up survey questionnaire. We then converted each option to numerical values as follows: “Already working on improvement” = 4, “Intending to improve” = 3, “No intention to improve” = 2, and “No improvement needed” = 1.

To determine the specificity of the intervention effects on readiness to change drinking habits, we also included readiness to change diet and smoking habits at 12 and 24 weeks as secondary outcomes. For readiness to change diet, each option was converted to numerical values as follows: “Already working on improvement” = 4, “Intending to improve” = 3, “No intention to improve” = 2, and “No improvement needed” = 1. For readiness to change smoking habits, among participants who were current smokers at baseline, the options were converted as follows: “Smoked but quit” = 3, “Intending to improve” = 2, and “No intention to improve” = 1.

#### Physician adherence with the Ultra-BI procedure

At cluster level, we asked physicians at the Ultra-BI clusters to mark the screening survey questionnaire for each patient to confirm they had provided an oral feedback message on the screening survey and an alcohol-information leaflet as our measure of physician adherence with the Ultra-BI procedure. Additionally, the follow-up survey questionnaire included a question asking participants, “Did you receive advice regarding your drinking habits from the physician during the consultation on the day you participated in the survey in July-August 2023?” We categorised the responses as follows: “Received” = participant answered “Received” at either the 12-week or 24-week follow-up; “Did not receive”, = participant answered, “Did not receive” at either the 12-week or 24-week follow-up or at any point; “Unsure” = did not answer “Received” or “Did not receive” at any time.

### Intervention and control

#### Ultra-BI group

In the intervention clinics, all eligible patients were screened with the Alcohol Use Disorders Identification Test (AUDIT). Those who met the aforementioned AUDIT-C inclusion criterion received the Ultra-BI. At the clinic reception, screening participants responded to the AUDIT and the questionnaire that included readiness to change their drinking behaviour. After the screening, the trial participants received an alcohol-information leaflet and brief oral message from their physician at the end of the consultation. The contents of the Ultra-BI were developed based on the materials and procedures used in our pilot trial in a general hospital setting, which demonstrated promising results [15]. We designed both the leaflet and brief oral message in accordance with the FRAMES (feedback, responsibility, advice, menu of options, empathy, and self-efficacy) approach [26].

The double-sided leaflet is in a simple and compact format, using easy-to-understand illustrations and colours. It provides advice on appropriate drinking levels and tips for reducing alcohol consumption. The contents are as follows: 1) an explanation of the “standard drink” unit used to measure alcohol consumption and a conversion table for various types of alcoholic beverages, 2) guidelines for drinking levels, health risks associated with excessive drinking, and the health benefits of reducing alcohol intake, 3) specific steps for reducing alcohol consumption, including setting goals, recording progress, and self-evaluation, and 4) tips for avoiding excessive drinking and an introduction to tools, including a website and a smartphone app.

The template of the oral message is as follows: “Mr./Ms. [Name], you might drink too much.” (Feedback); “I recommend you calculate your alcohol consumption on your own.” (Advice); “The information here (in the leaflet) will be beneficial for you.” (chance of effectiveness); “You can easily apply these recommendations (in the leaflet) starting today.” (Assurance of feasibility); and “I look forward to hearing your thoughts when we meet next time” (Motivation through commitment).

Physicians in the Ultra-BI clusters were given a paper outlining the procedure for conducting the Ultra-BI, along with a demonstration video.

#### Simplified assessment only (SAO) group

In the control clinics, to better align the control condition with routine clinical practice which typically does not involve systematic screening for drinking behaviour, we asked all eligible participants to fill in a simplified questionnaire. This simplified questionnaire included the same items used in the Ultra-BI group but excluded AUDIT questions 4 to 10 and the item to assess readiness to change drinking behaviour. We removed these items because previous studies have suggested that completing the AUDIT or similar assessments of drinking behaviour can lead to improvements in drinking behaviour [23,24].

After completing the screening questionnaire, participants submitted their questionnaire responses to the reception, and the receptionist stored the questionnaires in a box to be sent back to the research centre. Therefore, the physicians in the SAO clusters were unaware of the questionnaire responses and provided usual care to the participants.

### Financial incentives

Each cluster that enrolled at least one participant was given 50,000 JPY (approximately £245 GBP) to compensate for the staff time involved in executing the study procedures. Participants received a 500 JPY (approximately £2.45 GBP) gift card as a token of appreciation upon completing the questionnaires at baseline, as well as at the 12-week and 24-week follow-ups.

### Sample size

We set the sample size for the cluster RCT at 1,125 participants. This was determined based on an assumed effect size of 0.25 standardised mean difference for Ultra-BI, with a two-sided α level of 0.05, a power of 0.80, 40 clusters, and an intra-cluster correlation coefficient (ICC) between 0.03 and 0.04 [10]. Under these assumptions, we calculated the sample size necessary to detect the effect of Ultra-BI to be between 800 and 1,000 participants. Taking the median of this range, we estimated 900 participants for the cluster RCT. If we assume a dropout rate of 20% during the follow-up period of the RCT, the necessary sample size increased to 1,125 participants. To achieve the sample size, we instructed each cluster to conduct the screening survey until the number of participants with AUDIT-C scores of 5 or higher for men and 4 or higher for women reached a minimum of 29.

### Statistical analysis

We used SAS software version 9.4 (SAS Institute, Inc, Cary, NC, USA) and R version 4.2.3 [27]. The analysis included two populations: the intention-to-treat (ITT) population and the per-protocol (PP) population. The ITT population consisted of all participants, who were analysed in the group to which they had been randomised. The PP population included participants who were provided the Ultra-BI intervention based on the physician’s report in the Ultra-BI group and all participants in the SAO group.

The primary analysis was conducted on the ITT population, but the same analysis was also performed on the PP population. A general linear mixed-effects model was used to estimate the least square means of past 4 weeks TAC for each group at each time point, along with the point estimates and 95% confidence intervals for the differences between groups [28]. The dependent variable was the TAC. The fixed effects in the model included study group (Ultra-BI vs. SAO), time point (12 weeks, 24 weeks), baseline TAC converted from the AUDIT-C score, the group by time point interaction, and participant characteristics at screening (gender, quantified age group (converted to a continuous variable by assigning median values to each group), and AUDIT-C score. The model considered the cluster as a random effect (random intercept). An unstructured variance-covariance matrix was assumed for the correlation between time points for the dependent variable, and the Kenward-Roger method was used to calculate the degrees of freedom. We analysed observed cases only, without imputation for missing data, under the assumption that missing data occurred at random [29,30]. We also calculated Hedges’ g based on the between-group differences and pooled standard deviations at each follow-up time point.

We calculated a Bayes factor for TAC at 24 weeks on the ITT population. Bayes factors compare how well data support the alternative hypothesis over the null hypothesis. A Bayes factor greater than 1 suggests that the alternative hypothesis fits the data better than the null hypothesis. A Bayes factor less than 1 indicates that the null hypothesis fits the data better than the alternative hypothesis. A Bayes factor equal to 1 suggests that the data fit both hypotheses equally well. Jeffreys’ cut-off provides guidelines for interpreting Bayes factors [31]. A Bayes factor between 1/10 and 1/3 indicates moderate evidence against the alternative hypothesis. A Bayes factor less than 1/10 indicates strong evidence against the alternative hypothesis. To calculate a Bayes factor, we used the ttestBF function from the R BayesFactor package. This function is based on a t-test for independent two-group means. Though replicating the primary analysis condition was ideal, it was not feasible within the constraints of the package [32].

For the analysis of readiness to change drinking habits, we used a general linear mixed-effects model similar to TAC on the ITT population. The dependent variable was the quantified readiness to change drinking habits, and the independent variables included the group, time point, the interaction term between group and time point, gender, quantified age group, and baseline TAC converted from the AUDIT-C score. Baseline readiness to change drinking habits was not included as a covariate because it was not collected in the SAO group. Similar models were used for diet and smoking habits analyses, with their respective baseline readiness to change scores as covariates instead of TAC.

Subgroup analyses were performed for both TAC and readiness to change drinking habits on the ITT population. The subgrouping variables included AUDIT-C score category (4–6, 7–9, 10–12), representing equal thirds of the possible score range, gender (male, female), age group (20–30s, 40s, 50s, 60s, 70s), and visit type (routine appointments or not) collected at screening survey. We illustrated the results of the subgroup analyses using forest plots.

Physician-reported adherence with the Ultra-BI at screening, WHO DRL, readiness to change drinking habits, and participant-reported perception of receiving advice at follow-up were aggregated by the allocation group.

### Ethical considerations

To closely mimic real-world clinical settings, potential participants were asked to provide written consent for trial participation after reading the informed consent document, without receiving a formal oral explanation. Additionally, to minimise the risk of expectation bias, we only mentioned that the study was related to health-related lifestyles. The study protocol, including these procedures, was approved by the institutional review board of the Kurihama Medical and Addiction Center on May 22, 2023 (registration number: 423). The entire study adhered to the Declaration of Helsinki, and we obtained written informed consent from all participants.

### Patient and public involvement

This study did not involve patients in the development of the research question, choice of outcome measures, study design.

### Changes from the protocol and the statistical analysis plan

The statistical analysis plan (SAP) version 1.0 was finalized on March 20, 2024, before conducting any statistical analyses. This version included descriptive statistics for baseline characteristics, estimation of between-group differences in TAC and readiness to change drinking habits using general linear mixed-effects models, and their corresponding subgroup analyses. On 28 November, 2024, we revised the SAP to align with the Guidelines for the Content of Statistical Analysis Plans in Clinical Trials [33]. No additional analyses were performed after this revision.

Of note, among the outcomes analysed, only TAC was pre-specified in the protocol, SAP version 1.0, and the trial registry. Analyses of readiness to change drinking habits, as well as diet and smoking habits, were inadvertently omitted from both the protocol and trial registration and are therefore considered post-hoc analyses. Bayes factor analyses and categorical summaries of TAC and readiness to change drinking habits were performed as post-hoc analyses not specified in SAP version 1.0.

## Results

Figure 2 illustrates the flow of participants in this cluster randomised controlled trial. Recruitment occurred from 29 June to 7 August, 2023, across 43 clinics, with three clinics withdrawing due to lack of participant screening. In total, 3,537 participants from 40 clinics consented to participate, with 1,631 in the Ultra-BI group and 1,906 in the SAO group. After excluding those not meeting the hazardous drinking criteria of the AUDIT-C or with missing data in AUDIT-C items, 531 participants in the Ultra-BI group and 602 in the SAO group were eligible for follow-up. Among the Ultra-BI group, 528 (99%) received the intervention adhering to the protocol. At either 12 weeks, 24 weeks, or both, 473 (89%) participants in the Ultra-BI group and 535 (89%) in the SAO group responded, with 417 (79%) in the Ultra-BI group and 464 (77%) in the SAO group responding at 12 weeks, and 450 (85%) in the Ultra-BI group and 503 (84%) in the SAO group responding at 24 weeks.

**Figure 2.**
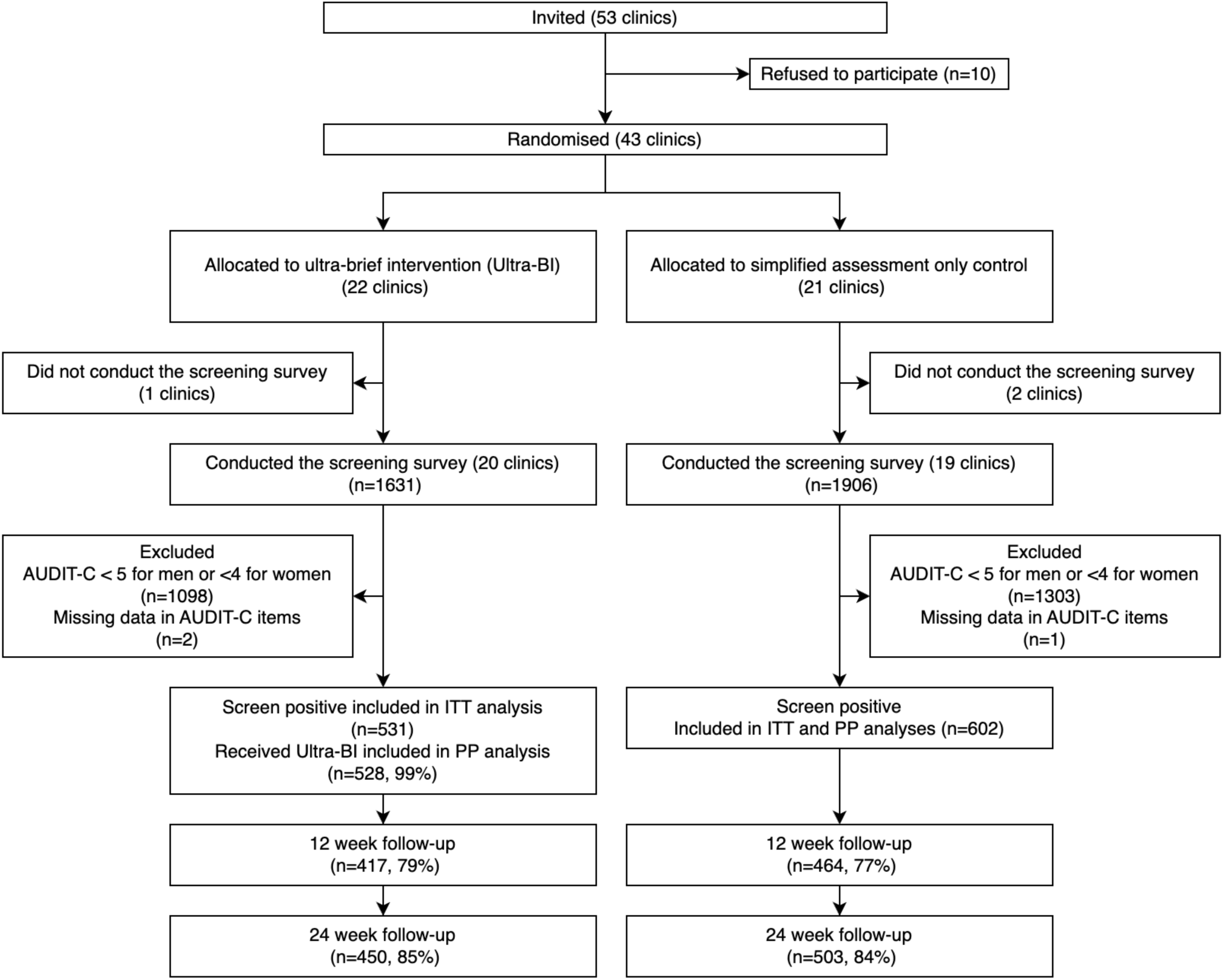
CONSORT flow diagram of clusters and patients through the trial Note: For the 12-week survey, we initially used registered mail requiring signature upon delivery; however, this resulted in some delivery failures due to recipient absence. Therefore, we switched to regular postal mail for the 24-week survey.

Table 1 presents the baseline characteristics of participants. Despite randomisation, the Ultra-BI group had a 5% higher proportion of female participants compared to the SAO group: 36% vs. 31%. Additionally, the mean TAC, calculated using AUDIT-C, was slightly higher in the Ultra-BI group compared to the SAO group: 749.9 g/4 weeks vs. 721.9 g/4 weeks.

**Table 1.** Baseline characteristics per participant by allocation group.

| Characteristics | Ultra-BI<br>n=531 | Simplified screening<br>assessment<br>n=602 | Total<br>n=1133 |
| --- | --- | --- | --- |
| Age group |  |  |  |
| 20–29 | 17 (3%) | 27 (4%) | 44 (4%) |
| 30–39 | 40 (8%) | 33 (5%) | 73 (6%) |
| 40–49 | 89 (17%) | 79 (13%) | 168 (15%) |
| 50–59 | 118 (22%) | 150 (25%) | 268 (24%) |
| 60–69 | 170 (32%) | 190 (32%) | 360 (32%) |
| 70–74 | 97 (18%) | 123 (20%) | 220 (19%) |
| % male | 342 (64%) | 417 (69%) | 759 (67%) |
| Visit type |  |  |  |
| First visit | 26/527 (5%) | 41/596 (7%) | 67/1123 (6%) |
| Routine appointments | 396/527 (75%) | 456/596 (77%) | 852/1123 (76%) |
| Visit as needed | 105/527 (20%) | 99/596 (17%) | 204/1123 (18%) |
| % smokers | 165/530 (31%) | 168/601 (28%) | 333/1131 (29%) |
| Comorbidity |  |  |  |
| Hypertension | 287 (54%) | 347 (58%) | 634 (56%) |
| Hyperuricemia | 74 (14%) | 62 (10%) | 136 (12%) |
| Diabetes | 75 (14%) | 88 (15%) | 163 (14%) |
| Dyslipidemia | 97 (18%) | 114 (19%) | 211 (19%) |
| Liver diseases | 60 (11%) | 63 (10%) | 123 (11%) |
| Digestive diseases | 88 (17%) | 105 (17%) | 193 (17%) |
| AUDIT-C score mean (SD) | 7.3 (2.4) | 7.2 (2.3) | 7.2 (2.3) |
| Baseline TAC per 4 weeks based<br>on AUDIT-C mean (SD) | 749.9 (525.5) | 721.9 (516.4) | 735.0 (520.1) |
| AUDIT-C=alcohol use disorders identification test - consumption, IQR=interquartile range, SD=standard deviation,<br>TAC=total alcohol consumption, Ultra-BI=ultra-brief intervention |  |  |  |
| Unless otherwise specified, the denominator for percentages is the total sample size shown at the top of each column.<br>For variables where the denominator differs, the actual denominator is indicated in that row. |  |  |  |

### Total alcohol consumption at follow-up

Table 2 and Figure 3 presents the TAC at 12 and 24 weeks for both the ITT and PP populations, showing least square means, group differences, and Hedge’s g estimated from general linear mixed-effects models. In the ITT population, the primary outcome of TAC at 24 weeks for the Ultra-BI group was 1046.9 g/4 weeks (95% CI 918.3 to 1175.4), and for the SAO group, it was 1019.0 g/4 weeks (95% CI 893.5 to 1144.6). The ICC for TAC was 0.04, as expected in the sample size calculations, which assumed a range of 0.03–0.04. The difference between groups was 27.8 g/4 weeks (95% CI -149.7 to 205.4), with a Hedge’s g of 0.02 (95% CI -0.10 to 0.14). The Bayes factor for the between-group difference was 0.08 ± 0.25, which, according to Jeffreys’ scale, indicates strong support for the null hypothesis of no difference between groups [31]. When categorising TAC by WHO drinking risk levels in the ITT population, 44% of the Ultra-BI group were classified as Abstinent or low risk, and 29% as Medium risk. In the SAO group, 46% were classified as Abstinent or low risk, and 29% as Medium risk (Table 3).

**Figure 3.**
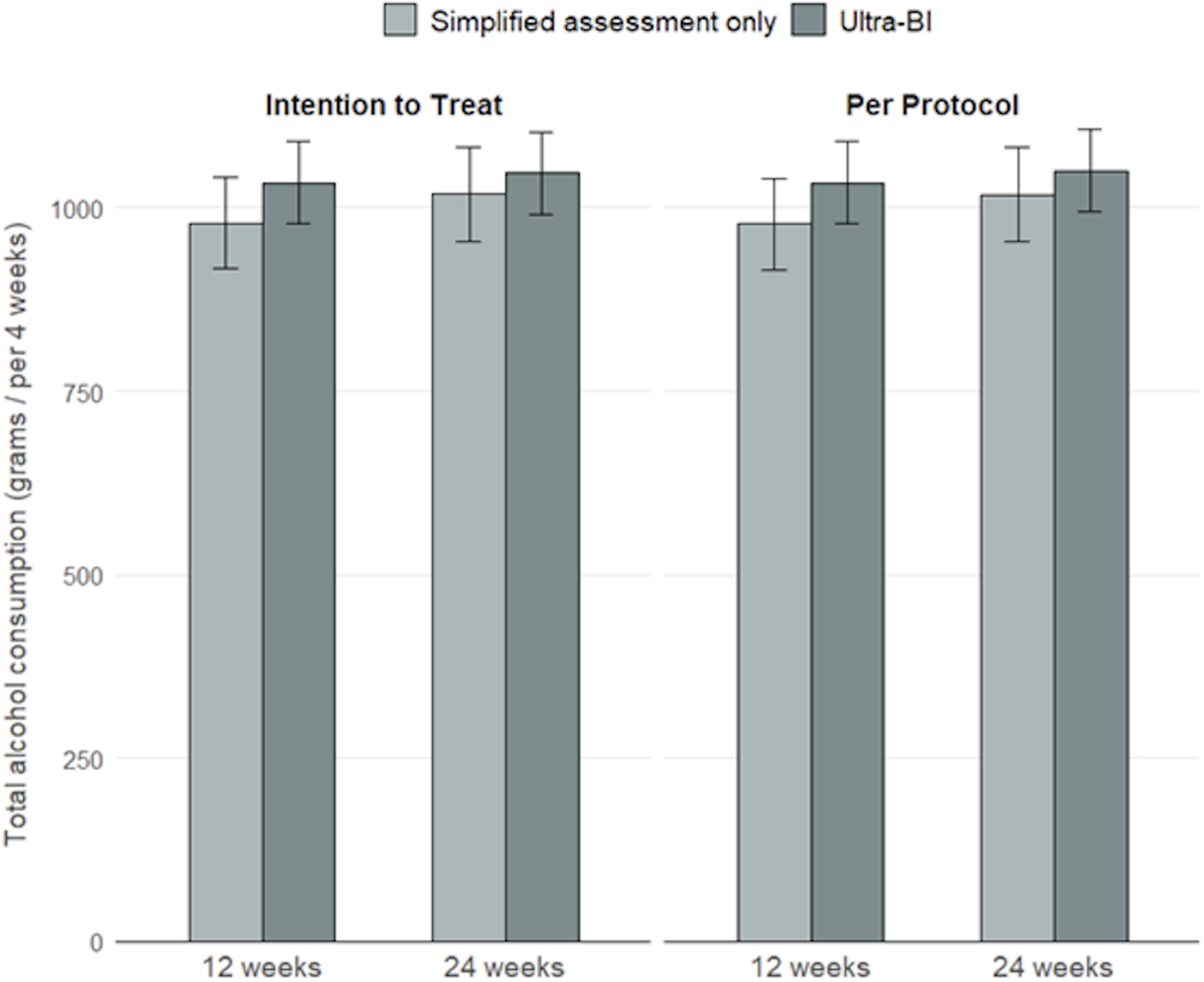
Total alcohol consumption in grams per 4 weeks at 12- and 24-week follow-up.

**Table 2.** Total alcohol consumption in grams per 4 weeks at 12 and 24 week follow-up.

| **Dataset** | **Time** | **Ultra-BI** | **Simplified assessment only** | **Difference (95% CI), P value** | **Hedge's g (95% CI)** |
| --- | --- | --- | --- | --- | --- |
| Intention to treat | 12 weeks | 1034.1<br>(919.6 to 1148.7) | 979.3<br>(866.1 to 1092.4) | 54.9<br>(-104.1 to 213.9), 0.49 | 0.04<br>(-0.08 to 0.16) |
|  | 24 weeks | 1046.9<br>(918.3 to 1175.4) | 1019.0<br>(893.5 to 1144.6) | 27.8<br>(-149.7 to 205.4), 0.75 | 0.02<br>(-0.10 to 0.14) |
| Per protocol | 12 weeks | 1034.4<br>(918.5 to 1150.3) | 978.0<br>(863.6 to 1092.4) | 56.4<br>(-104.4 to 217.2), 0.48 | 0.04<br>(-0.08 to 0.16) |
|  | 24 weeks | 1050.6<br>(920.9 to 1180.4) | 1017.8<br>(891.2 to 1144.5) | 32.8<br>(-146.5 to 212.1), 0.72 | 0.02<br>(-0.10 to 0.14) |

CI=confidence interval, Ultra-BI=ultra-brief intervention

**Table 3.** Proportion of WHO drinking risk level at follow-ups.

| Dataset | Group | Time | Abstinent or low risk | Medium risk | High risk | Very high risk |
| --- | --- | --- | --- | --- | --- | --- |
| Intention to treat | Ultra-BI | 12 weeks | 187/418 (45%) | 124/418 (30%) | 64/418 (15%) | 43/418 (10%) |
|  |  | 24 weeks | 198/453 (44%) | 133/453 (29%) | 78/453 (17%) | 44/453 (10%) |
|  | Simplified assessment only | 12 weeks | 216/465 (46%) | 141/465 (30%) | 66/465 (14%) | 42/465 (9%) |
|  |  | 24 weeks | 233/504 (46%) | 146/504 (29%) | 73/504 (14%) | 52/504 (10%) |

Ultra-BI=ultra-brief intervention

### Readiness to change drinking habits, diet, and smoking habits at follow-up

As shown in Table 4 and Figure 4, the readiness to change drinking habits, converted into numerical scores where higher scores indicate greater readiness to change, was higher in the Ultra-BI group compared to the SAO group at both 12 weeks (difference 0.30 [95% CI 0.10 to 0.40]; Hedge’s g 0.21 [95% CI 0.10 to 0.33]) and 24 weeks (difference 0.20 [95% CI 0.10 to 0.30]; Hedge’s g 0.16 [95% CI 0.05 to 0.28]). Additionally, at 12 weeks, 46% of the Ultra-BI group reported “Intending to improve” compared to 37% in the SAO group, and 16% versus 10% reported “Already working on improvement.” At 24 weeks, 44% of the Ultra-BI group and 41% of the SAO group reported “Intending to improve,” while 17% of the Ultra-BI group and 10% of the SAO group reported “Already working on improvement” (Table 5).

**Figure 4.**
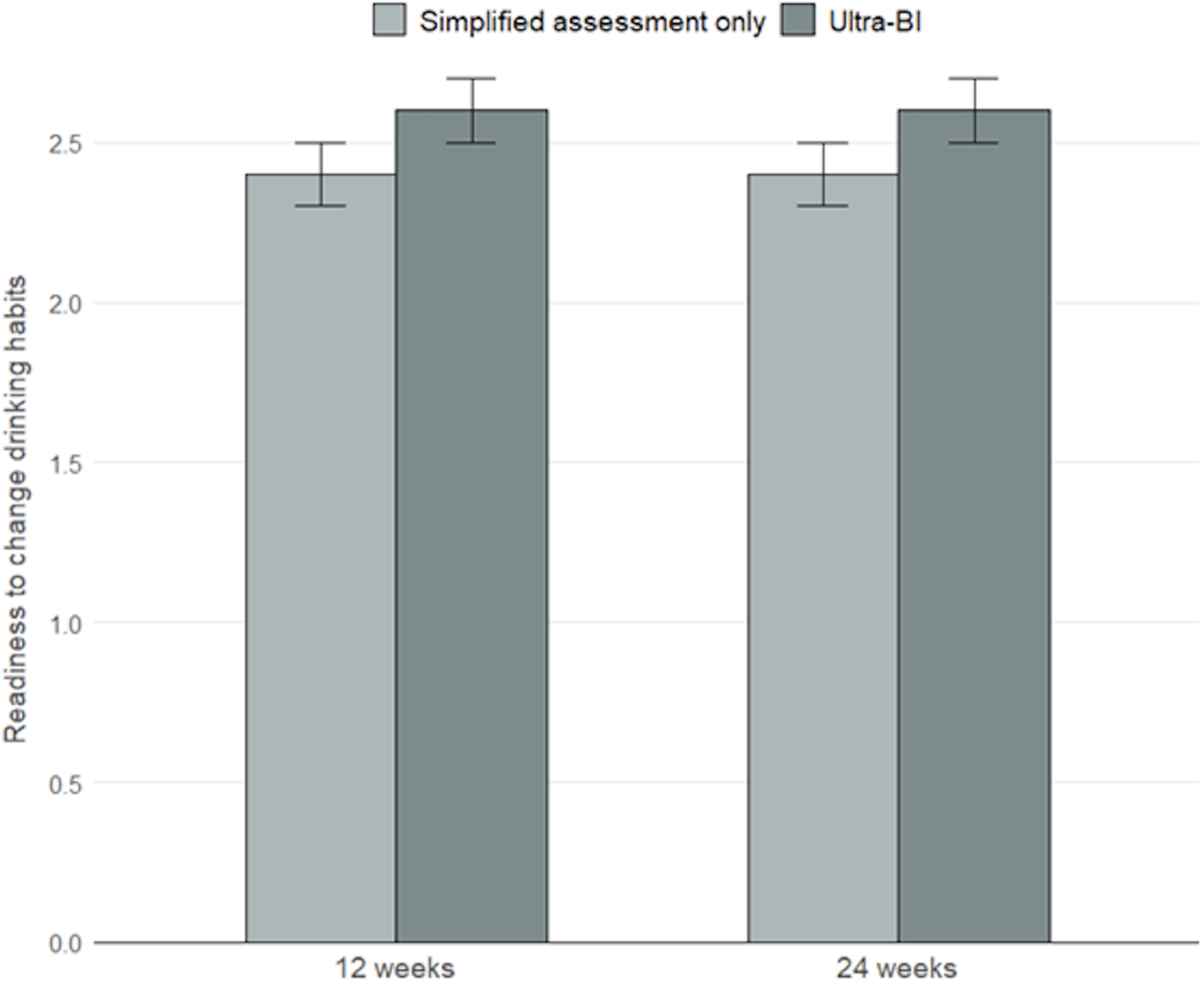
Readiness to change drinking habits at 12- and 24-week follow-up.

**Table 4.** Readiness to change drinking habits at 12- and 24-week follow-up.

| **Dataset** | **Time** | **Ultra-BI** | **Simplified assessment only** | **Difference (95% CI), P value** | **Hedge's g (95% CI)** |
| --- | --- | --- | --- | --- | --- |
| Intention to treat | 12 weeks | 2.6 (2.5 to 2.7) | 2.4 (2.3 to 2.5) | 0.3 (0.1 to 0.4), < 0.01 | 0.21 (0.10 to 0.33) |
|  | 24 weeks | 2.6 (2.5 to 2.7) | 2.4 (2.3 to 2.5) | 0.2 (0.1 to 0.3), < 0.01 | 0.16 (0.05 to 0.28) |

ICC=intraclass correlation coefficient, SE=standard error, Ultra-BI=ultra-brief intervention

**Table 5.**
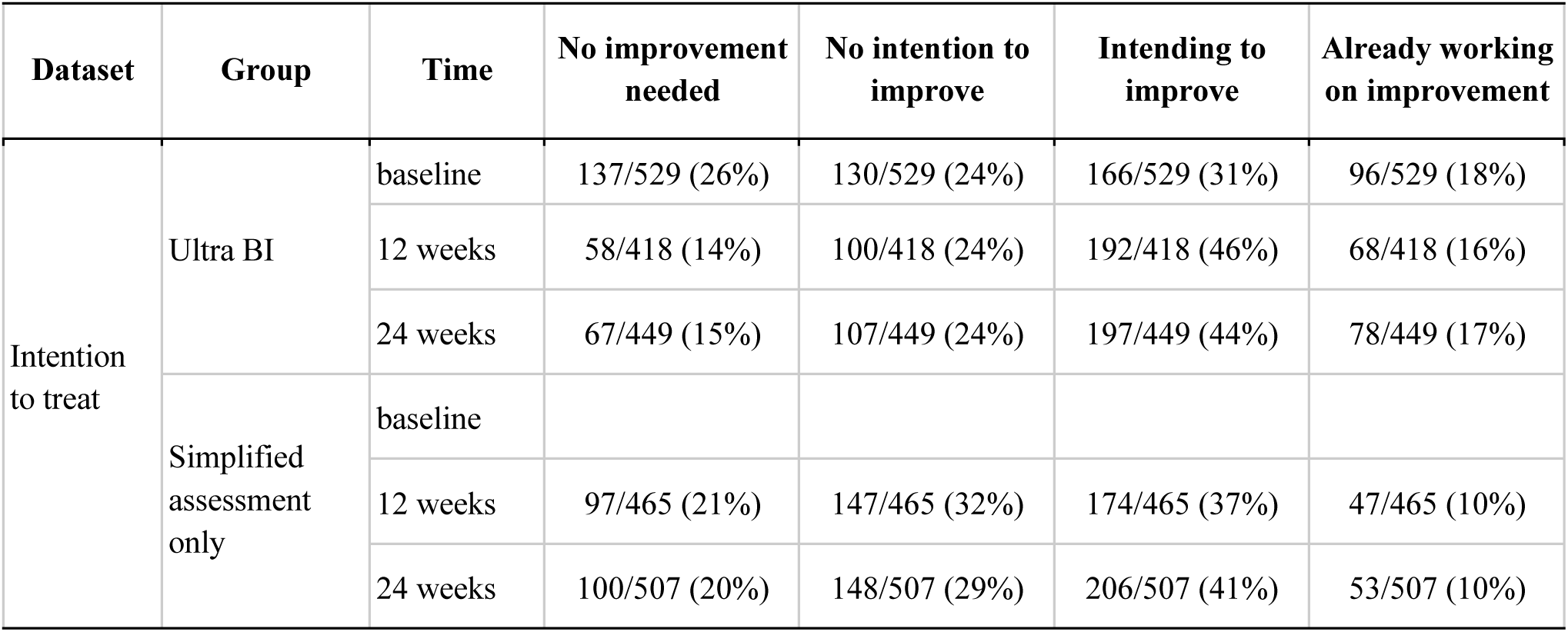
Proportion of readiness to change drinking habits by category.

Regarding readiness to change diet, the mean scores were 2.83 and 2.73 for the Ultra-BI and SAO groups at 12 weeks (difference 0.10 [95% CI -0.02 to 0.20]), and 2.89 and 2.86 at 24 weeks (difference 0.03 [95% CI -0.08 to 0.14]). For readiness to change smoking habits among baseline smokers, the mean scores were 1.75 and 1.73 at 12 weeks (difference 0.02 [95% CI -0.11 to 0.14]) and 1.79 and 1.86 at 24 weeks (difference -0.08 [95% CI -0.21 to 0.05]) for the Ultra-BI and SAO groups.

### Subgroup analysis

Figures 5 and 6 present the results of the subgroup analysis of the ITT population at 24 weeks for TAC and readiness to change drinking habits. The analysis found no evidence of interactions between any of the outcomes and the subgrouping variables.

**Figure 5.**
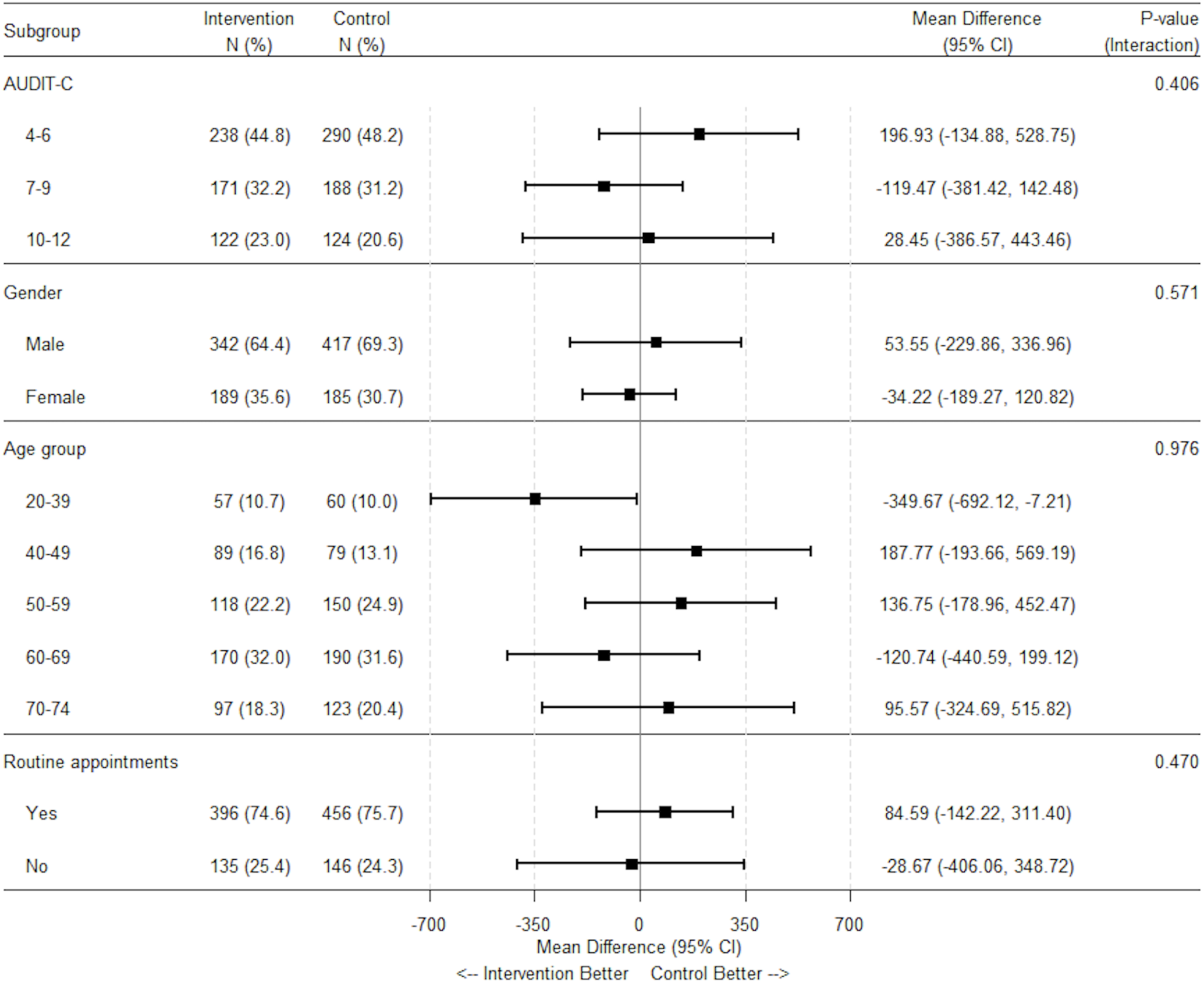
Subgroup analysis on total alcohol consumption per 4 weeks at 24-week follow-up.

**Figure 6.**
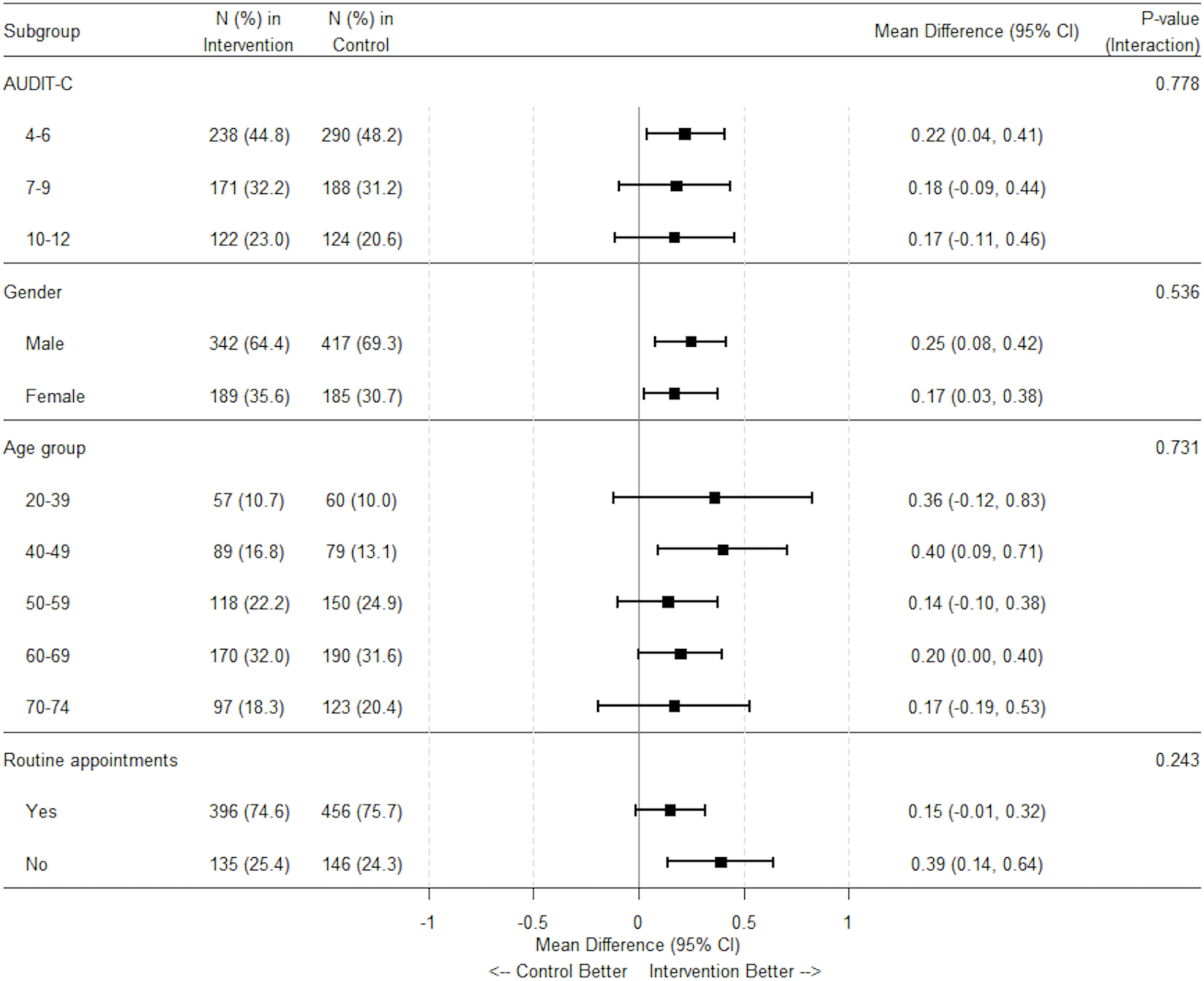
Subgroup analysis on readiness to change drinking habits at 24-week follow-up.

### Participant-reported perception of receiving advice

Of 475 participants in the Ultra-BI group, a total of 306 participants (64%) reported that they received advice from the physician, while 147 participants (31%) reported that they did not receive advice. Additionally, 22 participants (5%) reported that they were unsure whether they had received advice or not.

## Discussion

The present trial did not support the effectiveness of Ultra-BI on TAC at 12 and 24 weeks between the Ultra-BI and the SAO groups among patients engaging in hazardous drinking in Japanese primary care settings. The 95% CI for the difference in TAC at 24 weeks had an upper limit of 205.4 grams of alcohol per 4 weeks, corresponding to a Hedge’s g of 0.14. This suggests a trivial clinical benefit. Furthermore, the Bayes factor of 0.08 strongly supported the null hypothesis of no difference between the two arms. On the other hand, readiness to change drinking habits was more favourable in the Ultra-BI group at both 12 and 24 weeks. The absence of similar improvements in readiness to change diet or smoking habits suggests Ultra-BI specifically affected readiness to change drinking habits. These findings challenge current recommendations that advocate for the widespread implementation of BIs in primary care settings, while also highlighting potential benefits in affecting patient readiness to change that may be particularly relevant in the context of continuous care.

### Possible reasons for null findings

Contrary to our expectation, our results did not support the effectiveness of the Ultra-BI in primary care settings. Our pilot trial, which investigated the same Ultra-BI of this trial in general hospital wards, had shown promising results [15].The SIPS trial [10] in UK primary care settings had suggested that an Ultra-BI was as effective as 5 or 20-minute BIs. These findings led us to anticipate the effectiveness of Ultra-BIs in primary care. However, assuming our findings are correct, questions may arise about the effectiveness of both Ultra-BIs and BIs compared to assessment-only control in reducing alcohol consumption with a Japanese population. Alternatively, it may also mean that the Japanese population requires different motivational considerations in future BI program iterations. However, the possibility that our results are false negatives due to systematic or random errors should be considered.

Systematic errors could arise from participants, training intensity, intervention adherence, and control conditions. The effectiveness of Ultra-BI may be diminished in participants with higher severity. There is limited evidence supporting the effectiveness of BIs for people with dependence or very heavy drinking [34]. However, our subgroup analysis did not provide evidence of effect modification for TAC by the AUDIT-C score category.

The low intensity of the training for physicians, which involved only watching a video, might have reduced the effect of Ultra-BI due to insufficient physician confidence or passion in addressing alcohol-related issues. However, the lack of time among healthcare professionals is often cited as a barrier to the implementation of BIs [9]. Therefore, even if more intensive training could improve the intervention’s effect, it might also reduce its broad implementation, leading to a lesser public health impact.

The possibility that the intervention was not delivered as intended has been suggested as a reason for the null findings of trials [35]. Although physicians reported delivering Ultra-BI to most participants, only two-thirds of those who responded to follow-up surveys in the Ultra-BI group perceived that they had received advice from their physicians, indicating this potential issue. However, the proportion of participants who perceived they had received advice might be considered an indicator of the intensity of Ultra-BI rather than that of physicians’ nonadherence. Even in a pilot study conducted in a general hospital, where the researcher who developed Ultra-BI administered the intervention to all participants and demonstrated promising results, only 70% of participants in the intervention group reported that they were given feedback by the physician [15]. Given these proportions, it is likely that many participants who perceived they had not received advice did receive it. However, in this non-treatment seeking population, the fact that Ultra-BI improved readiness to change drinking habits represents a meaningful step toward behaviour change, even though it did not immediately lead to reduced alcohol consumption.

Too strong control conditions can lead to false negative findings in RCTs of BIs. Even screening itself may have some effect over no treatment at all, and mask the effect of the BIs being tested [23,24]. However, we made the screening survey questionnaire for the control SAO group as simple as possible. If the intervention cannot demonstrate superiority over this control, its effect can be considered not reaching the level of clinical importance.

Random error may have resulted in a greater baseline TAC in the Ultra-BI group, potentially leading to false negative results. The baseline TAC, converted from AUDIT-C scores, was approximately 30g/4 weeks higher in the Ultra-BI group compared to the SAO group. Additionally, the AUDIT-C has a ceiling effect when measuring TAC. The frequency of drinking option “≥ 4 days per week” converts to 22 days per 4 weeks, and the typical quantity option “≥ 10 drinks” converts to 100g. Therefore, the actual baseline TAC difference between groups might be larger than indicated by AUDIT-C scores. However, we included the baseline TAC, converted from AUDIT-C scores, as a covariate in the efficacy analysis, accounting for this difference. Furthermore, subgroup analysis did not suggest the superiority of the Ultra-BI group in any category including the highest AUDIT-C score category, where the ceiling effect would have had the most impact on the results.

### Strengths and limitations of the trial

The trial has several strengths, including a large sample size (N=1,133), high follow-up rates (84%), and the control condition close to real-world “no intervention” settings by simplifying the study procedures and questionnaires. The use of Bayesian analysis allowed for assessing the evidence for no effect, beyond simply failing to find evidence of an effect. Additionally, the narrow 95% CI of Hedge’s g for our primary outcome suggests that any true effectiveness of the Ultra-BI is trivial, even at best. Furthermore, this trial contributes valuable evidence on BI in primary care settings among Asian populations, where such data have been notably scarce. Previous systematic reviews of BI in these settings included only one trial from an Asian country, with Asians comprising merely 1% of participants in Western studies [8]. These strengths contribute to a nuanced understanding of the intervention’s potential clinical impact in real-world settings.

However, some limitations should be acknowledged. Baseline TAC and readiness to change drinking habits were not collected. Although this was to avoid the potential effect of intensive screening, balancing between-group differences in these outcomes at baseline due to random error became difficult. Reliance on self-reported invitation and participation rates from clusters, as well as their self-reported adherence with the intervention protocol without direct observation, may have introduced bias towards no effect. Yet, strict adherence monitoring could differ from daily practice. Furthermore, the findings are specific to general screening and Ultra-BIs in primary care settings in Japan. Studies indicate BI effectiveness can vary across ethnicities [36]. Readers should exercise caution when interpreting our findings in an international context.

## Implications

Despite these limitations, the trial has important implications for alcohol screening and brief interventions in primary care. From a public health perspective, our results suggest that widely implementing Ultra-BI in primary care may not lead to clinically relevant benefits to reduce alcohol consumption. However, from a clinical standpoint, some physicians may recognize the importance of Ultra-BI based on its impact on readiness to change, given that the strength of primary care lies in continuity of care. Further pragmatic effectiveness trials of other forms of BIs including those harnessing information technologies and artificial intelligence within the constraints of primary care may be warranted [37].

In conclusion, we found evidence suggesting no to minimal benefit at best by the Ultra-BI compared to simplified assessment only in reducing drinking among patients with hazardous drinking or suspected alcohol dependence in Japanese primary care settings. Our findings challenge the validity of current recommendations for screening and brief interventions in primary care.

## Funding

The EASY study was conducted with a grant from the Japan Agency for Medical Research and Development (AMED) (23he0122018j0003), awarded to the consortium comprising CureApp, Inc., the Kurihama Medical and Addiction Center, and Okayama City Hospital. AMED had no involvement in any aspect of the study process.

## Data Availability

All data produced in the present study are available upon reasonable request to the authors.

## Acknowledgments

We thank all staff members who contributed to conducting this study at each clinic. Additionally, we would like to express gratitude to NEO MARKETING INC. for their efficient management of the logistics for the EASY study.

## Declaration of Generative AI and AI-assisted technologies in the writing process

During the preparation of this manuscript, we used chatGPT, Claude, and Grammarly for proofreading and sentence revision of the English language but not for their intellectual content. After using these tools, we thoroughly reviewed and edited the content where necessary. We assume full responsibility for the content of the publication.

---

| **Author name** | **CRediT authorship contribution statement** |
| --- | --- |
| Ryuhei So | Conceptualization, Methodology, Formal analysis, Data Curation, Writing - Original Draft, Writing - Review & Editing, Project administration, Funding acquisition |
| Kazuya Kariyama | Conceptualization, Methodology, Writing - Review & Editing |
| Shunsuke Oyamada | Methodology, Formal analysis, Resources, Data Curation, Writing - Review & Editing |
| Sachio Matsushita | Conceptualization, Methodology, Writing - Review & Editing, Funding acquisition |
| Hiroki Nishimura | Visualization, Writing - Review & Editing |
| Yukio Tezuka | Methodology, Resources, Writing - Review & Editing |
| Takashi Sunami | Methodology, Resources, Writing - Review & Editing |
| Toshi A. Furukawa | Methodology, Writing - Review & Editing, Supervision |
| Ethan Sahker | Writing - Review & Editing |
| Mitsuhiko Kawaguchi | Investigation, Writing - Review & Editing |
| Haruhiko Kobashi | Writing - Review & Editing |
| Sohji Nishina | Investigation, Writing - Review & Editing |
| Yuki Otsuka | Writing - Review & Editing |
| Yasushi Tsujimoto | Investigation, Writing - Review & Editing |
| Yoshinori Horie | Conceptualization, Writing - Review & Editing |
| Hitoshi Yoshiji | Conceptualization, Writing - Review & Editing |
| Takefumi Yuzuriha | Conceptualization, Writing - Review & Editing |
| Kazuhiro Nouse | Conceptualization, Methodology, Writing - Review & Editing, Funding acquisition |

**sTable 1.** Baseline characteristics per cluster by allocation group.

| Characteristics | Ultra-BI | Simplified assessment only | Total |
| --- | --- | --- | --- |
| Per cluster: | n=21 | n=19 | n=40 |
| Median (IQR) N of participants | 29 (21–32) | 30 (29–34) | 29 (28–32) |
| Median (IQR) Age group |  |  |  |
| 20–29 | 0% (0–6%) | 2% (0–5%) | 1% (0–6%) |
| 30–39 | 7% (3–10%) | 3% (3–7%) | 4% (3%–10%) |
| 40–49 | 16% (10–22%) | 13% (8–15%) | 14% (9–20%) |
| 50–59 | 19% (14–33%) | 23% (17–28%) | 20% (16–32%) |
| 60–69 | 30% (26–39%) | 31% (24–38%) | 31% (25–38%) |
| 70–74 | 17% (7–26%) | 23% (9–26%) | 20% ( 8–26%) |
| Median (IQR) % male | 66% (53–72%) | 71% (62–78%) | 68% (58–76%) |
| Median (IQR) AUDIT-C score | 7.2 (6.7–7.6) | 7.0 (6.9–7.3) | 7.1 (6.8–7.6) |
| Median (IQR) Baseline TAC per 4 weeks based on AUDIT-C | 731.0 (665.5–787.8) | 715.7 (643.2–765.4) | 719.8 (662.7–773.1) |
| IQR=interquartile range, SD=standard deviation, Ultra-BI=ultra-brief intervention |  |  |  |
| Unless otherwise specified, the denominator for percentages is the total sample size shown at the top of each column. For variables where the denominator differs, the actual denominator is indicated in that row. |  |  |  |

## Notes

### Competing Interest Statement

Ryuhei So and Hiroki Nishimura were employed by CureApp, Inc. during the conduct of the study. Ryuhei So received institutional grants from the Osake-no-Kagaku Foundation and the Mental Health Okamoto Memorial Foundation. He holds multiple pending patents (JP2022049590A, US20220084673A1, JP2022178215A, JP2022070086, JP2023074128A). Sachio Matsushita received personal fees from CureApp, Inc., Otsuka Pharmaceutical Co., Ltd., EA Pharma Co., Ltd., Takeda Pharmaceutical Co., Ltd., Yoshitomi Pharmaceutical Industries, Ltd., Eisai Co., Ltd., Nippon Shinyaku Co., Ltd., and MSD K.K. outside the submitted work. Toshi A. Furukawa reports personal fees from Boehringer-Ingelheim, Daiichi Sankyo, DT Axis, Micron, Shionogi, SONY and UpToDate, and a grant from DT Axis and Shionogi, outside the submitted work; In addition, Toshi A. Furukawa has a patent 7448125 and a pending patent 2022-082495, and has licensed intellectual properties for Kokoro-app to Mitsubishi-Tanabe. Yukio Tezuka received lecture fees from MSD K.K. outside the submitted work. Yoshinori Horie, Hitoshi Yoshiji received personal fees from CureApp, Inc. and Otsuka Pharmaceutical Co., Ltd., outside the submitted work. Takefumi Yuzuriha received personal fees from CureApp, Inc., Otsuka Pharmaceutical Co., Ltd., and Nippon Shinyaku Co., Ltd., outside the submitted work. Kazuhiro Nouso received personal fees from CureApp, Inc. for coordinating a pivotal study outside the submitted work. All other authors declare no competing interests.

### Clinical Trial

UMIN000051388

### Author Declarations

The study protocol, including these procedures, was approved by the institutional review board of the Kurihama Medical and Addiction Center on May 22, 2023 (registration number: 423).

## References

1 World Health Organization. Global Status Report on Alcohol and Health 2018. World Health Organization 2019.

2 Reid MC, Fiellin DA, O’Connor PG. Hazardous and harmful alcohol consumption in primary care. Arch Intern Med. 1999;159:1681–9.

3 Gache P, Michaud P, Landry U, et al. The Alcohol Use Disorders Identification Test (AUDIT) as a screening tool for excessive drinking in primary care: reliability and validity of a French version. Alcohol Clin Exp Res. 2005;29:2001–7.

4 So R, Kariyama K, Oyamada S, et al. Prevalence of hazardous drinking and suspected alcohol dependence in Japanese primary care settings. Gen Hosp Psychiatry. 2024;89:8–15.

5 Babor TF, Higgins-Biddle JC, Saunders JB, et al. The Alcohol Use Disorders Identification Test: Guidelines for use in primary care. World Health Organization. 2001.

6 Kuitunen-Paul S, Roerecke M. Alcohol Use Disorders Identification Test (AUDIT) and mortality risk: a systematic review and meta-analysis. J Epidemiol Community Health. 2018;72:856–63.

7 Nilsen P, Aalto M, Bendtsen P, et al. Effectiveness of strategies to implement brief alcohol intervention in primary healthcare. A systematic review. Scand J Prim Health Care. 2006;24:5– 15.

8 Kaner EF, Beyer FR, Muirhead C, et al. Effectiveness of brief alcohol interventions in primary care populations. Cochrane Database Syst Rev. 2018;2:CD004148.

9 Johnson M, Jackson R, Guillaume L, et al. Barriers and facilitators to implementing screening and brief intervention for alcohol misuse: a systematic review of qualitative evidence. J Public Health . 2011;33:412–21.

10 Kaner E, Bland M, Cassidy P, et al. Effectiveness of screening and brief alcohol intervention in primary care (SIPS trial): pragmatic cluster randomised controlled trial. BMJ. 2013;346:e8501.

11 Dienes Z, Coulton S, Heather N. Using Bayes factors to evaluate evidence for no effect: examples from the SIPS project. Addiction. 2018;113:240–6.

12 Heather N. Interpreting null findings from trials of alcohol brief interventions. Front Psychiatry. 2014;5:85.

13 Holloway AS, Watson HE, Arthur AJ, et al. The effect of brief interventions on alcohol consumption among heavy drinkers in a general hospital setting. Addiction. 2007;102:1762–70.

14 Freyer-Adam J, Baumann S, Haberecht K, et al. In-person alcohol counseling versus computer-generated feedback: Results from a randomized controlled trial. Health Psychol. 2018;37:70–80.

15 Tezuka Y, So R, Fukuda T. The effectiveness of an ultra-brief intervention in 1 min for hazardous drinking in a general hospital setting: A quasi-randomized pilot trial. Psychiatry and Clinical Neurosciences Reports. 2024;3:e216.

16 Wang TC, Kyriacou DN, Wolf MS. Effects of an intervention brochure on emergency department patients’ safe alcohol use and knowledge. J Emerg Med. 2010;39:561–8.

17 Cunningham JA, Neighbors C, Wild C, et al. Ultra-brief intervention for problem drinkers: results from a randomized controlled trial. PLoS One. 2012;7:e48003.

18 Campbell MK, Piaggio G, Elbourne DR, et al. Consort 2010 statement: extension to cluster randomised trials. BMJ. 2012;345:e5661.

19 Saunders JB, Aasland OG, Babor TF, et al. Development of the Alcohol Use Disorders Identification Test (AUDIT): WHO collaborative project on early detection of persons with harmful alcohol consumption--II. Addiction. 1993;88:791–804.

20 Ikeda N, Takimoto H, Imai S, et al. Data Resource Profile: The Japan National Health and Nutrition Survey (NHNS). Int J Epidemiol. 2015;44:1842–9.

21 Bradley KA, DeBenedetti AF, Volk RJ, et al. AUDIT-C as a brief screen for alcohol misuse in primary care. Alcohol Clin Exp Res. 2007;31:1208–17.

22 Osaki Y, Ino A, Matsushita S, et al. Reliability and validity of the alcohol use disorders identification test—consumption in screening for adults with alcohol use disorders and risky drinking in Japan. Asian Pac J Cancer Prev. 2014;15:6571–4.

23 McCambridge J, Day M. Randomized controlled trial of the effects of completing the Alcohol Use Disorders Identification Test questionnaire on self-reported hazardous drinking. Addiction. 2008;103:241–8.

24 Kypri K, Langley JD, Saunders JB, et al. Assessment may conceal therapeutic benefit: findings from a randomized controlled trial for hazardous drinking. Addiction. 2007;102:62–70.

25 Desapriya E, Stockwell T, Doll SR, et al. International guide for monitoring alcohol consumption and related harm. Published Online First: 2000. doi: 10.13140/RG.2.2.29007.48808

26 Miller W, Sánchez V. Motivating young adults for treatment and lifestyle change. Alcohol use and misuse by young adults. 1994;198:55–81.

27 R Core Team. R: A Language and Environment for Statistical Computing. 2022.

28 Bell ML, Rabe BA. The mixed model for repeated measures for cluster randomized trials: a simulation study investigating bias and type I error with missing continuous data. Trials. 2020;21:148.

29 Taljaard M, Donner A, Klar N. Imputation strategies for missing continuous outcomes in cluster randomized trials. Biom J. 2008;50:329–45.

30 Hossain A, Diaz-Ordaz K, Bartlett JW. Missing continuous outcomes under covariate dependent missingness in cluster randomised trials. Stat Methods Med Res. 2017;26:1543–62.

31 Kass RE, Raftery AE. Bayes Factors. J Am Stat Assoc. 1995;90:773–95.

32 Morey R, Rouder J. BayesFactor version 0.9.9: An R package for computing Bayes factor for a variety of psychological research designs. Published Online First: 10 July 2014.

33 Gamble C, Krishan A, Stocken D, et al. Guidelines for the content of statistical analysis plans in clinical trials. JAMA. 2017;318:2337–43.

34 Saitz R. Alcohol screening and brief intervention in primary care: Absence of evidence for efficacy in people with dependence or very heavy drinking. Drug Alcohol Rev. 2010;29:631–40.

35 Heather N. The efficacy-effectiveness distinction in trials of alcohol brief intervention. Addict Sci Clin Pract. 2014;9:13.

36 Field CA, Caetano R, Harris TR, et al. Ethnic differences in drinking outcomes following a brief alcohol intervention in the trauma care setting: Ethnicity and brief intervention. Addiction. 2010;105:62–73.

37 Kaner EFS, Beyer FR, Garnett C, et al. Personalised digital interventions for reducing hazardous and harmful alcohol consumption in community-dwelling populations. Cochrane Database Syst Rev. Published Online First: 2017. doi: 10.1002/14651858.CD011479.pub2

